# COMPOUNDING ASSOCIATIONS OF EDUCATION AND SOCIAL CARE SUPPORT ON HOSPITAL COSTS THROUGHOUT CHILDHOOD

**DOI:** 10.64898/2026.08.11.26360173

**Authors:** Yiu-Shing Lau, Ruth Gilbert, German Pulido, Matt Sutton

## Abstract

**Objective:** To describe variation in hospital costs among children with different combinations of health conditions, special educational needs or disability (SEND) and children social care (CSC) indicators.

**Study Setting and Design:** This cross-sectional study used regression analysis to test whether two-way and three-way interactions of cross-public sector service use (health, education and social care) are associated with higher hospital costs in England.

**Data Sources and Analytic Sample:** Hospital care costs between April 2022 and March 2023 for the 8.9 million children aged 5-18 years were obtained from linked administrative hospital, education or social care data in the ECHILD database. Children were classified into eight categories based on combinations of indicators of chronic health conditions, SEND or CSC.

**Principal Findings:** Over one-third (35.4%) of children had some hospital costs during the year. Average costs were £317 for all children and £895 for children with non-zero hospital costs. By age 18, few children had no indicator in any sector (35.1% of boys, 43.7% of girls) and indicators in all three sectors were not rare (7.1% of boys, 6.2% of girls). At age 5, children with indicators recorded in all three sectors had the highest hospital costs (£2,952 for boys and £3,674 for girls). At age 18, males and females with indicators in all three sectors accounted for 21% and 23% of hospital costs, respectively. SEND and social care indicators without chronic health conditions were associated with only slightly higher hospital costs. Hospital costs were much higher for children with SEND if they also had a chronic health condition. Hospital costs were only higher for children with social care if they also had both a chronic health condition and SEND.

**Conclusions.:** Taking account of additional support from non-health sectors is important for understanding health sector costs. The compounding associations between use of other public sectors on health sector costs indicates scope for targeting of integrated care.

## Introduction

There is growing demand for hospital services in many high income countries which is applying pressure on the delivery and affordability of health care systems ^1^. Increases in population age structures are usually mentioned as a reason for increases in hospital demand, however, there is also evidence of increased demand for paediatric healthcare and healthcare costs ^2^.

Many studies have shown how children with chronic health conditions have higher hospital care costs than other children, especially those with multiple chronic health conditions or who require additional aids ^3–5^. The increasing complexity of child health needs is expected to place considerable pressure on health and related public services. Children with chronic conditions, disabilities, or social care involvement often require coordinated support, but children with such multiple needs are not reliably coded in healthcare data. Therefore, there is no comprehensive study of these additional health care costs. The ECHILD database linkage of administrative healthcare, education and social care, enables assessment of contacts in all three services for all children in England.

Previous studies have shown that children with special educational or social care support experience poorer health outcomes ^6–8^. However, few studies have examined how education ^9^ and social care support ^10^ interact with chronic health conditions to influence healthcare costs. Moreover, there is no evidence on how the cost implications of these complex higher needs vary across childhood.

We describe variation in hospital costs among children with different combinations of health conditions or support indicators measured in three public sectors. We use indicators from hospital, education or social care records of a chronic health condition or provision of support for SEND or for social care. We use linked national administrative data on 8.9 million children in 2022/23 to offer a national picture of how additional support in schools for SEND, or for social care, in combination with indicators of chronic health conditions recorded in hospital data, influence hospital costs for children aged 5 to 18 years in England.

## Methods

### Data sources

We use the Education and Child Health Insights from Linked Data (ECHILD) database version 2 ^11,12^, which is a linked administrative dataset in England that combines health, education, and social care data for children and young people. Education data is obtained from the National Pupil Database ^13^ consisting of school censuses in spring, summer and autumn for each academic year, which contains termly records indicating support for SEND. Information on support from social care services comes from local authority returns on Children in Need (CiN: e.g. children with contact with children social care excluding cases with no further action) ^14^ and Children Looked After (CLA, children in care) ^13^. Hospital data is obtained from Hospital Episode Statistics (HES) on hospital admissions ^15^ (including critical care), and outpatient visits ^16^. We also use the Emergency Care Dataset (ECDS) ^17^ to source attendances at emergency departments.

### Population

We used the spring, autumn and summer school censuses of the 2022/23 academic year from the National Pupil Database (NPD) to identify children living in England aged five to 16 years as at the 1^st^ of April 2022. Children aged 17 and 18 at the 1^st^ of April 2022 were obtained from the spring, summer and autumn censuses of the 2021/22 and 2020/21 academic years, respectively. In total our population included 8,869,800 children who were aged between five and eighteen in these extracts of the NPD.

To reduce the probability that the same child would be duplicated because they were given a separate unique pupil number, we omitted children in Alternative Provision and Pupil Referral Unit censuses as well as children that were in nursery national curriculum years (“N1” and “N2”). To further minimise duplication of children, we kept only records with two types of enrolment status: “C = Current (single registration at this school)” and “M = Current Main (dual registration)”. Thus, we excluded records with the following enrolment status: “G = Guest (pupil not registered at this school but attending some lessons or sessions)”, “S = Current Subsidiary (dual registration)”, “F = FE College” and “O = Other provider”. Our population is defined only by the NPD, so we did not include children who were identified only through a birth record or hospital records. Information on the identification of the population base is provided in Appendix A table A1.

### Costing

We calculated hospital costs for all children in the population incurred during the financial year 2022/23 (April to March) using activity recorded in Hospital Episode Statistics and the Emergency Care Dataset. We followed the costing approach used by NHS England in the estimation of the resource allocation formula for General and Acute hospital services ^18^. We did not include costs occurred by mental health service providers as we do not have access to a fill financial year of mental health services activity within ECHILDS v2.

The costing involves a three-step iterative process to apply costs to all Admitted Patient Care and Outpatient appointments. The costing procedure uses Healthcare Resource Group (HRG) codes ^19,20^ which are standard groupings of clinically similar treatments that use similar levels of healthcare resources. First, we applied NHS tariffs ^21^ to all activity that could be attached to an HRG. Tariffs for each HRG vary by admission mode (whether planned or unplanned) and by the length of stay (with a per diem rate added for long hospital spells as directed by NHS tariffs). Not all activity has an NHS tariff price, particularly high-cost items such as drugs, devices or procedures. An example is procedures relating to soft tissue sarcomas. These high-cost activities may be rare in nature and as such not suitable for national prices and are usually locally agreed. For activity that could not be costed using the NHS Tariffs, we used NHS Reference Costs ^22^. These unit costs also vary by length of stay (for admissions only: crude differences which are long or short stay with no per diem cost) and admission mode. Activity may not have a reference cost or tariff price if the hospital did not provide sufficient information to link the activity to any HRG, so in a third step, we applied an average cost by specialty to all remaining activity. We provide the proportion of total costs estimated by each costing procedure in appendix B.

Critical care data within ECHILDs Version 2 does not include HRG codes. Instead, we used admission and discharge dates to generate length of stay in critical care and critical care unit function codes to determine the type of paediatric and neonatal care. Critical care activity was costed using the per diem costs for each of the critical care functions using NHS Reference Costs.

Emergency care activity was costed using national tariffs for the 12 relevant HRG codes ^23^. The HRG assigned to emergency care activity is mainly dependent on the investigations and treatments undertaken. Separate categories are applied for dental care and patients that are recorded as dead on arrival. In 2022/23, the tariffs varied between £87 to £420 depending on the type of care provided.

To reduce the impact of approximately 80 individuals that had extremely high costs, we truncated the annual costs for individual children at £500,000. Further details on the contributions of each costing procedure are provided in Appendix B.

### Indicators of health conditions or additional support

For health conditions, we used diagnostic codes in any hospital admission record relating to a chronic health condition (CHC) (Hardelid et al. 2014) recorded at any point from birth up to the end of March 2023. Chronic health conditions in children are defined as any health problem more than 50% likely to require follow-up more than one year after the event is recorded in a hospital admission ^25^. This information is extracted using Admitted Patient Care data from HES for financial years 2004/5 to 2022/23 using International Classification of Diseases version 10. These include birth records for all children born in the NHS in England. Mental health data was not used to code mental health CHCs as diagnostic coding is generally incomplete.

Additional educational support is termed special educational needs or disability (SEND). This was derived using information on whether a child had ever had a record for low intensity support funded by the school, called SEN support or the more intensive Education and Health Care Plan (EHCP), funded by the local authority ^26^. This information was extracted from the autumn, spring and summer censuses for academic years from 2003/4 until 2022/23. Special educational provision is educational or training provision that is additional to or different from that made generally for other children or young people of the same age in mainstream education ^27^. One third of children in state education have ever had a record of SEND provision by age 16 ^28^.

Social care support was derived using information on whether a child had ever had a record in CiN or CLA returns. If a child had been referred to Children in Need but that referral resulted in no further action, then we did not classify that referral as indicating social care support. Children who use social care are assessed as being in need, may be subject to child protection plans, may have disabilities and may be looked after. We extracted CiN data for all children in our sample from 2008/09 academic year and CLA from 2003/4 until 2022/23 academic years. We were unable to extract CiN data prior to 2008/09 as this was when the data was first made available. This means for children aged 13 and older, social care status represents only CLA.

The indicators for the three sectors are not mutually exclusive. Some children have indicators from two of the three sectors and some have indicators from all three sectors. We classify individual children into eight categories representing: one category with no indicator from any sector; three categories with an indicator from one sector only; three categories with indicators from two sectors; and one category with indicators from all three sectors.

### Analysis

We assigned all children to one of the eight categories and calculated the contribution of each category to the total costs for this population by combinations of year-of-age and gender. We then summarised the average costs of children in each of the eight needs categories by combinations of year-of-age and gender.

We used linear regression to test whether two-way and three-way interactions between the indicators for the three sectors were associated with significant additional impacts on costs. The constant term in the regression applies to all observations and gives the estimated average costs for children who have no needs indicators from any sector. The three main associations (CSC, SEND and CHC) show by how much average costs are increased if a child has this needs indicator present. For example, the average costs for a child with a CHC and no needs indicated in other sectors equals the regression constant plus the estimated main CHC association. The coefficients on the three two-way interactions (SEND & CHC, CSC & CHC and CSC & SEND) show the additional costs, over and above the sum of the two associated main associations, for a child who has needs indicators from the two sectors. For example, the average costs for a child with CHC and SEND equals the regression constant, plus the main association of CHC, plus the main association of SEND, plus the additional association from the two-way interaction of CHC and SEND. The coefficient on the three-way interaction represents any further additional association on hospital costs of having all three needs indicators. The average cost for a child in this needs category equals the sum of all eight coefficients. The statistical significance of the three two-way interactions and the single three-way interaction are the tests for whether there are compounding associations of needs from multiple sectors.

We perform two sets of regression models for sensitivity. Firstly, we redefined the indicators relating to SEND and CSC. We change SEND to represent only children who ever had an Education and Health Care Plan and CSC to include only CLA children. Secondly, we only classify children as SEND if they had SEND support recorded in the four years before 2022/23. We limit the observation period for SEND identification in the years prior to 2022/23 to capture more recent SEND support, in the main analysis children could have required SEND support in their early ages and then stopped requiring support throughout their teenage lives.

All analyses were conducted using Stata 18 MP4.

## Results

### Prevalence of each needs category by gender and age

At the age of 18 years, only 37% of males and 46% of females had had no needs indicator recorded in any of the three sectors. Conversely, 6.3% of males and 5.5% of females had needs indicators from all three sectors by the age of 18 years. Prevalence rates of CHC only remain stable at around 9% for males and increase from 8.4% to 11.6% for females. We find that the rates of children with CHC increase with age for both genders, but these increases are also linked to having CHC and other need indicators. Summing across categories in Table 1, 21.7% of males have had SEND only, however 48% of individuals have either SEND, SEND & CSC and SEND & CHC, making males with SEND the largest category. Females with SEND are a smaller proportion of the population at 14% at the age of 18 (32% with SEND, SEND & CSC, SEND & CHC). At the age of 18, there is little difference in the percentages of males and females that have a history of social care needs, however, there are a higher percentage of females with CSC only.

**Table 1:** Prevalence of each of the eight categories by age and gender.

|  | None | CSC | SEND | CHC | CSC &<br>SEND | CSC &<br>CHC | SEND &<br>CHC | ALL |
| --- | --- | --- | --- | --- | --- | --- | --- | --- |
| Age | Males |  |  |  |  |  |  |  |
| 5 | 69.07 | 1.39 | 13.60 | 9.77 | 0.83 | 0.26 | 4.72 | 0.37 |
| 6 | 64.15 | 2.43 | 15.79 | 9.17 | 1.82 | 0.47 | 5.40 | 0.77 |
| 7 | 60.02 | 3.28 | 16.99 | 9.05 | 2.82 | 0.64 | 5.97 | 1.24 |
| 8 | 56.80 | 3.80 | 18.12 | 8.82 | 3.68 | 0.75 | 6.29 | 1.73 |
| 9 | 53.87 | 4.38 | 18.70 | 8.91 | 4.45 | 0.90 | 6.69 | 2.10 |
| 10 | 51.61 | 4.82 | 18.84 | 8.98 | 5.21 | 1.04 | 6.93 | 2.57 |
| 11 | 49.13 | 5.06 | 19.02 | 9.19 | 5.94 | 1.22 | 7.35 | 3.10 |
| 12 | 46.84 | 5.38 | 19.35 | 9.17 | 6.76 | 1.36 | 7.49 | 3.64 |
| 13 | 45.06 | 5.59 | 19.67 | 9.08 | 7.46 | 1.42 | 7.62 | 4.10 |
| 14 | 43.99 | 5.61 | 19.75 | 9.05 | 8.07 | 1.48 | 7.59 | 4.47 |
| 15 | 42.35 | 5.73 | 20.09 | 8.92 | 8.64 | 1.61 | 7.72 | 4.93 |
| 16 | 40.34 | 5.67 | 20.54 | 9.03 | 9.17 | 1.69 | 8.06 | 5.51 |
| 17 | 38.49 | 5.44 | 21.02 | 9.16 | 9.64 | 1.75 | 8.39 | 6.09 |
| 18 | 37.43 | 5.08 | 21.70 | 9.40 | 9.37 | 1.73 | 8.99 | 6.31 |
|  | Females |  |  |  |  |  |  |  |
| 5 | 80.21 | 1.85 | 6.44 | 8.41 | 0.44 | 0.26 | 2.23 | 0.17 |
| 6 | 75.91 | 3.31 | 7.89 | 8.27 | 1.04 | 0.48 | 2.69 | 0.40 |
| 7 | 72.12 | 4.49 | 9.18 | 8.28 | 1.67 | 0.65 | 2.93 | 0.68 |
| 8 | 68.72 | 5.49 | 10.10 | 8.37 | 2.33 | 0.84 | 3.25 | 0.90 |
| 9 | 66.18 | 6.18 | 10.82 | 8.46 | 2.80 | 1.01 | 3.40 | 1.15 |
| 10 | 63.54 | 6.91 | 11.26 | 8.70 | 3.39 | 1.20 | 3.58 | 1.40 |
| 11 | 60.59 | 7.58 | 11.68 | 9.17 | 3.91 | 1.45 | 3.91 | 1.71 |
| 12 | 57.76 | 8.03 | 11.98 | 9.49 | 4.57 | 1.76 | 4.25 | 2.18 |
| 13 | 55.54 | 8.24 | 12.20 | 9.86 | 5.15 | 1.99 | 4.42 | 2.61 |
| 14 | 53.53 | 8.33 | 12.48 | 10.13 | 5.43 | 2.26 | 4.73 | 3.10 |
| 15 | 51.45 | 8.35 | 12.83 | 10.22 | 5.97 | 2.53 | 4.99 | 3.66 |
| 16 | 49.03 | 8.31 | 13.22 | 10.42 | 6.38 | 2.90 | 5.40 | 4.34 |
| 17 | 47.08 | 8.01 | 13.56 | 10.82 | 6.60 | 3.23 | 5.62 | 5.08 |
| 18 | 45.79 | 7.20 | 13.96 | 11.60 | 6.23 | 3.58 | 6.11 | 5.54 |

Notes: Percentages are row percentages for each age and gender. CSC represents children who have received social care support. SEND represents children with a special educational need or disability or children with an education of health care plan. CHC represents children with a Hardelid chronic health condition. ALL represents children with CSC, SEND and CHC.

**Table 2:** Numbers of children, proportions with non-zero hospital costs and average hospital costs, by gender and age.

| Age | Male |  |  | Female |  |  |
| --- | --- | --- | --- | --- | --- | --- |
|  | % With Costs | Total | Average cost (£) | % With Costs | Total | Average cost (£) |
| 5 | 45 | 324,270 | 375 | 39 | 308,240 | 306 |
| 6 | 40 | 331,390 | 313 | 36 | 313,790 | 279 |
| 7 | 37 | 329,000 | 288 | 33 | 312,910 | 252 |
| 8 | 36 | 330,850 | 280 | 32 | 315,230 | 245 |
| 9 | 36 | 341,830 | 283 | 32 | 325,400 | 245 |
| 10 | 36 | 344,010 | 285 | 32 | 328,280 | 243 |
| 11 | 37 | 340,240 | 317 | 32 | 323,120 | 263 |
| 12 | 37 | 332,730 | 310 | 33 | 317,000 | 296 |
| 13 | 37 | 327,910 | 329 | 34 | 314,290 | 350 |
| 14 | 37 | 325,080 | 344 | 36 | 309,320 | 381 |
| 15 | 35 | 311,820 | 344 | 36 | 297,640 | 397 |
| 16 | 32 | 304,720 | 325 | 35 | 290,990 | 401 |
| 17 | 32 | 302,710 | 317 | 37 | 288,820 | 427 |
| 18 | 31 | 292,780 | 286 | 39 | 279,820 | 447 |

Notes: Counts are rounded to the nearest 10 to abide by disclosure rules set by Office of National Statistics Secure Research Service.

### Average costs by gender and age

During 2022/23, 3,140,160 (35.4%) of the 8,864,160 children had some hospital costs. The mean costs for children with some hospital activity was £895 and the mean cost for all children was £317.

Average costs of hospital services vary by age and gender with the highest average costs incurred at age five for males (£375) and age 18 for females (£447) ( (32%) and increasing proportions throughout the teenage years.

For females, average costs are lowest at the age of 10 (£243). For males, average costs are lowest at eight years of age (£280). For males, costs increase during the start to the mid teenage years before reducing when reaching adulthood. For females, average costs reduce from birth to the age of 10 and increase throughout the teenage years.

### Total population costs associated with children in each category by gender and age

The proportions of total population costs represented by children in each of the eight categories are shown in figure 1. As the proportion of children with high needs increases, the proportion of total population costs increases. At age 18, males with indicators in all three sectors account for 21% of total hospital costs and females account for 23% of total hospital costs, despite being only 6% and 7% of the population, respectively. Conversely, at age eight years, males with no indicators contributed 16% of total costs and females with no indicators contributed 15% of total costs, despite being 35% and 44% of the population respectively.

**Figure 1:**
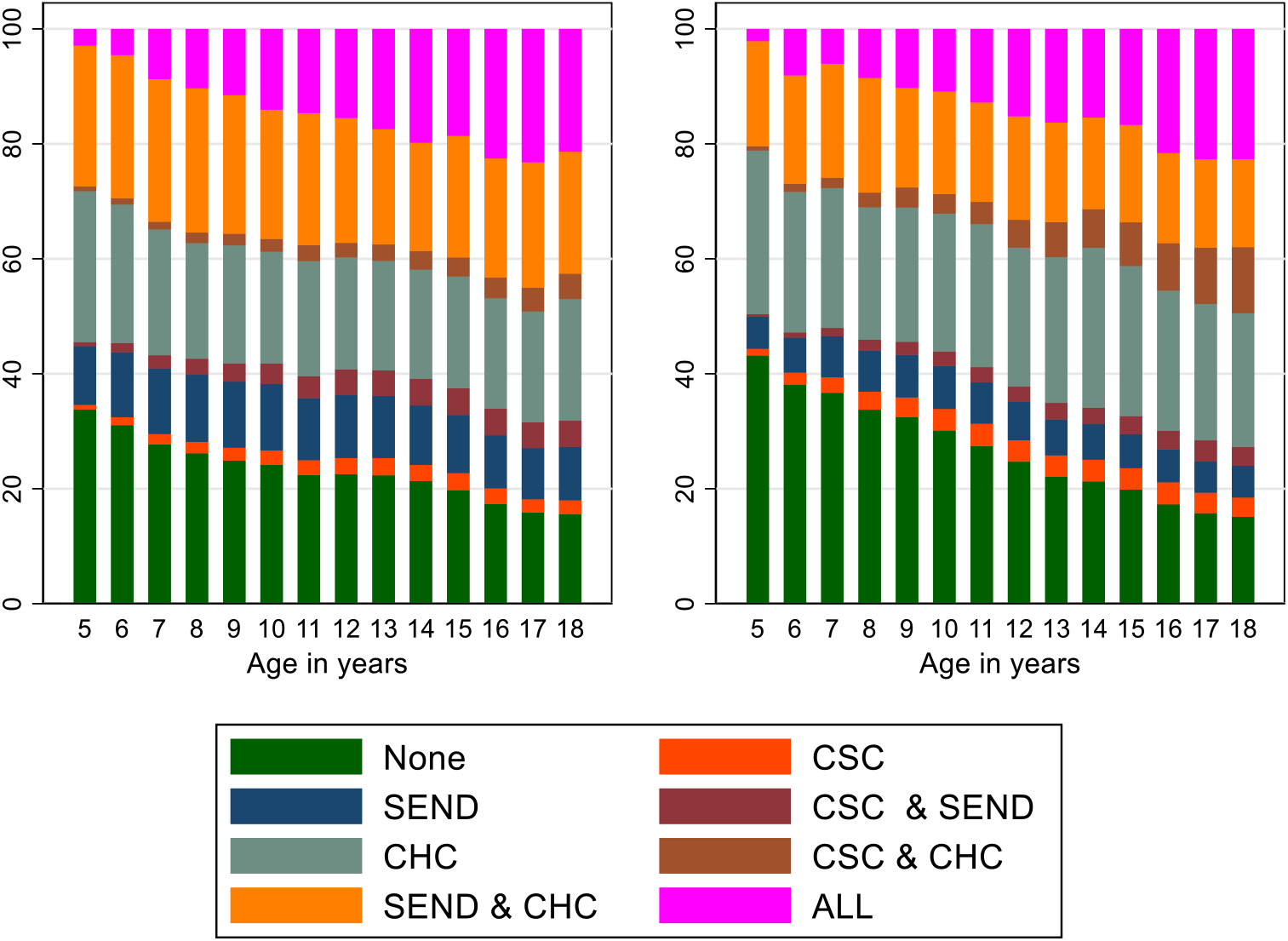
Proportions of total population costs associated with children in each of eight categories by age and gender. Notes: Left panel are costs for males and right panel are costs for females. CSC represents children who have received social care support. SEND represents children with a special educational need or disability or children with an education of health care plan. CHC represents children with a Hardelid chronic health condition. ALL represents children with CSC, SEND and CHC.

The highest percentages of children with non-zero hospital costs are also at ages five for males (45%) and 18 for females (39%). The proportion of males having non-zero hospital costs decreases as age increases, to 31% at age 18. We observe a different pattern for females, with the lowest proportion with non-zero hospital costs at 10 years old (32%) and increasing proportions throughout the teenage years.

For females, average costs are lowest at the age of 10 (£243). For males, average costs are lowest at eight years of age (£280). For males, costs increase during the start to the mid teenage years before reducing when reaching adulthood. For females, average costs reduce from birth to the age of 10 and increase throughout the teenage years.

### Total population costs associated with children in each category by gender and age

The proportions of total population costs represented by children in each of the eight categories are shown in figure 1. As the proportion of children with high needs increases, the proportion of total population costs increases. At age 18, males with indicators in all three sectors account for 21% of total hospital costs and females account for 23% of total hospital costs, despite being only 6% and 7% of the population, respectively. Conversely, at age eight years, males with no indicators contributed 16% of total costs and females with no indicators contributed 15% of total costs, despite being 35% and 44% of the population respectively.

### Average costs for each needs category by gender and age

Figure 2 shows average costs in each of the eight needs categories by gender and age. Boys and girls with needs indicators from all three sectors have the highest hospital costs across all ages. At almost all ages, children with CHC and SEND are the group with the second highest average costs. Children with CHC and CSC have similar costs to those with CHC only at most ages, except girls above the age of 12. Average costs across the population are highest for children aged five and reduce up to age 18. Children with CHC and CHC & SEND have higher costs in the teenage years.

**Figure 2:**
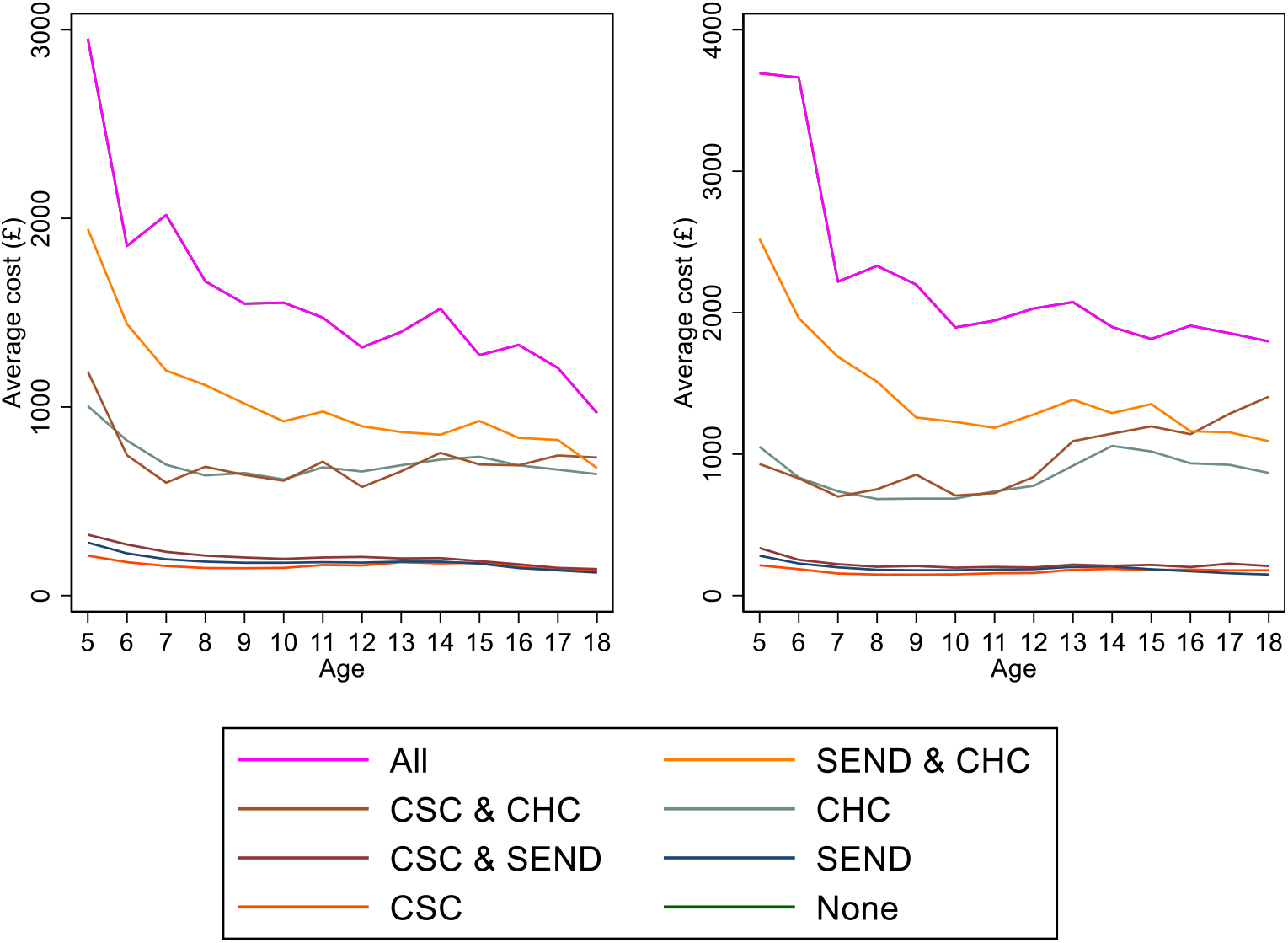
Average costs of the population in each needs category by age and gender.

The numbers underlying the Figure are provided in appendix B table B2. A figure only for children without CHCs is also provided in appendix B figure B1.

### Testing whether there are interactions between the associations of needs from different sectors

Figure 1 suggests that the indicators from the three sectors may have interacting associations on hospital costs. In this section we present the results from the regression modelling that formally tests the significance of those interactions. We estimated models for each combination of year-of-age and gender and Figure 3 shows the estimated coefficients for six of the age-specific models. We include the results for ages 5, 7, 10, 12, 15 and 18 years. Full regression results for all ages are included in Appendix C.

**Figure 3:**
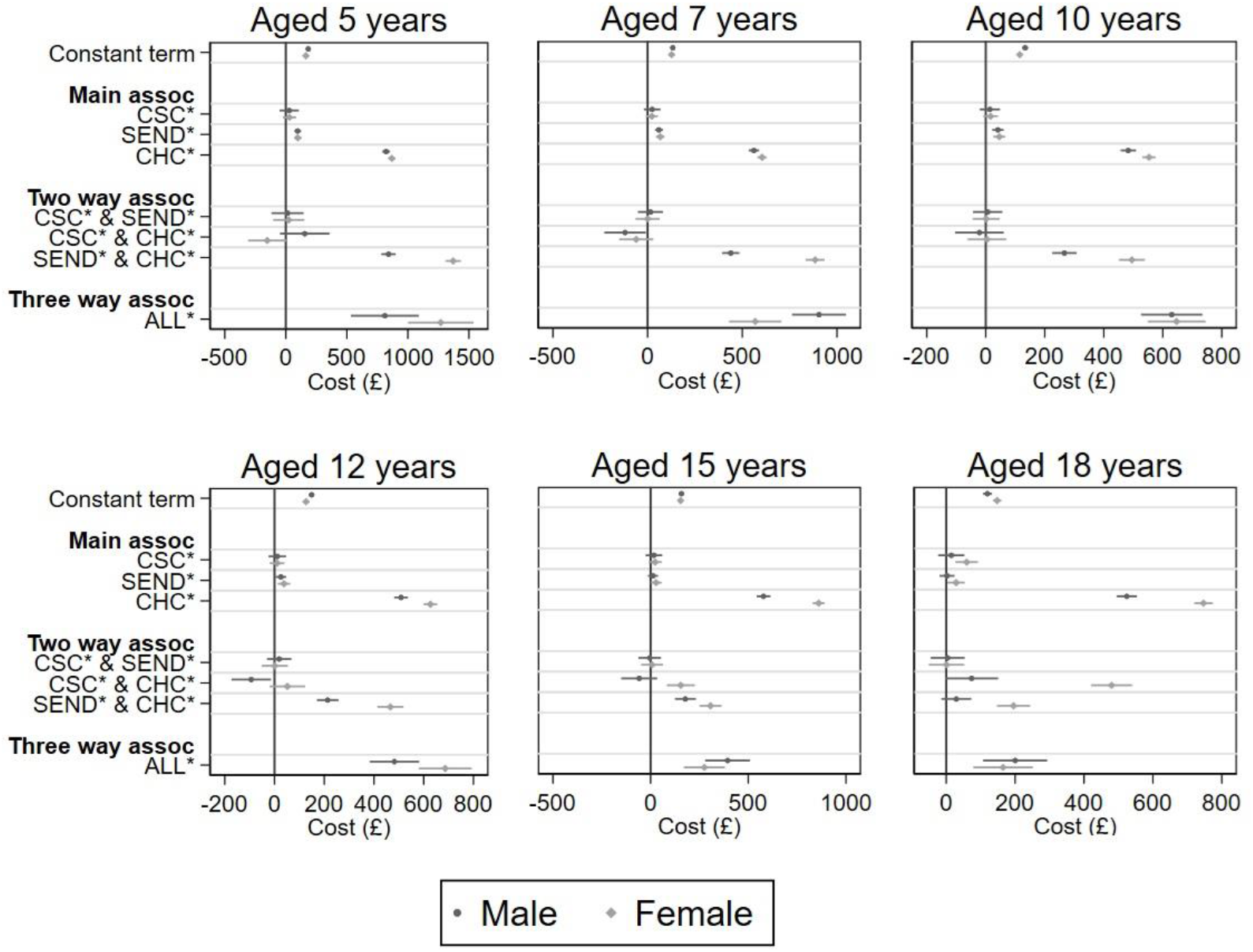
Estimated main and interaction associations at ages 5, 7, 10, 12, 15 and 18 years. Notes: Assoc is an abbreviations for associations. Coefficients are from age and gender stratified linear regressions including all eight terms (constant term, three main associations, three two-way interactions, and one three-way interaction). 95% confidence intervals shown around each estimate. CSC is an indicator for whether the child has received social care support. SEND is an indicator for whether the child has a special educational need or disability or an educational health care plan. CHC is an indicator for whether the child has a chronic health condition. ALL represents children with CSC, SEND and CHC.

At all ages (except 18 years for boys), the combination of CHC and SEND creates strong compounding associations on hospital costs. Other two-way interactions are not significant, except for SEND and CSC amongst girls aged 15 years and over.

Despite representing relatively small numbers of children in some cases, there is clear evidence of compounding associations of needs in three sectors for both genders at all ages.

### Sensitivity analysis

After redefining the SEND and CSC indicators to include only children with the higher levels of support in these sectors, we find similar results to the main analyses. The costs associated with only SEND and CSC increase because children with these indicators also have chronic health conditions (Appendix C table C2). We find that CSC is associated with having higher costs when compared to individuals without CSC, but CSC does not have any additional impact on costs when combined with any other indicator (CHC or SEND). In this supplementary analysis we do not find a significant additional association on costs for children with all three indicators, but this may be due to the low number of children in this category.

Models where we restrict SEND to being children who had SEND recorded in the prior 4 years produced similar results to the main analysis (Appendix C: tables C5 and C6). We find that the costs of individuals with only SEND remained similar to the main models, however, the additional costs of having SEND and at least one other indicator increased.

## Discussion

### Main messages

The 6% of children with indicators from all three sectors accounted for 21% of total hospital costs at age 18. The main predictor of hospital costs was CHCs. The 26% of children that have a CHC, with or without support indicators, accounted for 70% of hospital costs at age 18. SEND alone was associated with increased hospital costs, but this reduced for children in their teenage years. We found no associations of CSC alone with hospital costs, except for females aged 18 years.

SEND has compounding associations when combined with CHC. CSC has compounding associations on hospital costs only if it accompanied both SEND and CHC. These results provide evidence to support the need for integrated strategies between health care, education and children social care.

There are differences in the association of CHC on hospital utilisation for males and females. The differences in the main association of CHC for males and females increases over time. This is also reflected in compound associations CHC & SEND and also CHC & CSC. The gender differences may be due to the types of chronic conditions that males and females develop over time.

### Strengths

This is the first study at the population level that links hospital costs and indicators of needs from both children’s social care and special educational needs. Using ECHILD, we could link records at an individual level to all inpatient, outpatient, emergency department and critical care records to the National Pupil Database, which enables the linkage of a CSC use and SEND via ECHILD. We could thereby associate hospital costs with need indicators from other public services.

### Limitations

This study suffers from several limitations. Firstly, we focus on children that are recorded in State education between 5 and 18 years in the NPD. Children from more affluent backgrounds are under-represented because NPD only includes State schools (see Appendix A table A1). Secondly, there may be administrative linkage errors and approximately 2.2% of people aged 0-19 had opted out of the national health data by July 2023 ^29^. Nevertheless, we have been able to link 96.9% of the NPD population to a hospital record. Thirdly, we relied on indicators from English data and children who were diagnosed with a CHC or received education or social care support elsewhere would not be flagged in our data. Therefore, we under-represent the true proportions of children with additional needs. We estimated that we underrepresented the prevalence of CHC by 1% when children are aged five and by 3% when individuals are aged 18 years (see appendix A Table A3). Fourthly, we focused on school-age children which would have the possibility of requiring SEND and due to data linkages, CSC. This excludes the years of age when hospital costs are highest. Fifthly, we defined the indicators by whether they ever appeared in a child’s history. This may under-estimate the association of having more recent SEND or CSC support on hospital costs. Finally, we focus only on acute hospital costs and do not consider mental health, community health or primary care services.

## Conclusion

We find that needs recorded in other public sectors have compounding associations on hospital costs, particularly SEND. This provides some evidence that service integration may lead to reduction in hospital service utilisation. However, there is nuance in the findings, as males and females do not exhibit similar relationships.

## Acknowledgements

SL and GP were funded by the National Institute for Health Research (NIHR) Children and Families Policy Research Unit (NIHR206114). ECHILD is supported by ADR UK (Administrative Data Research UK), an Economic and Social Research Council (part of UK Research and Innovation) programme (ES/V000977/1, ES/X003663/1, ES/X000427/1).

## Ethics statement

Permissions to use linked, de-identified data from HES and the NPD from the ECHILD database were granted by NHS Digital (DARS-NIC-381972-Q5F0V-v0.5) and DfE (DR200604.02B). Patient consent was not required to use the deidentified data in this study.

## Data availability statement

The ECHILD database is made available for free for approved research based in the UK, via the ONS Secure Research Service. Enquiries to access the ECHILD database can be made by emailing. Researchers will need to be approved and submit a successful application to the ECHILD Data Access Committee and ONS Research Accreditation Panel to access the data, with strict statistical disclosure controls of all outputs of analyses.

This work contains statistical data from ONS which is Crown Copyright. The use of the ONS statistical data in this work does not imply the endorsement of the ONS in relation to the interpretation or analysis of the statistical data. This work uses research datasets which may not exactly reproduce National Statistics aggregates.

## Appendix A: Children in population

We obtained a population of children from the NPD that are aged between five and eighteen years of age on the 1^st^ of April 2022. Using spring, summer and autumn censuses of the National Pupil Database, we obtain a population of children from a school census closest to 1^st^ April 2022. Figure A1 shows a population flow diagram of the numbers of children in the population, along with the number of children which were dropped due to stated reasons. As mentioned in the main text, we do not include children that were in national curriculum years N1 and N2 (nursey), as well as Alternative Provision and Pupil Referral Unit censuses.

**Figure A1:**
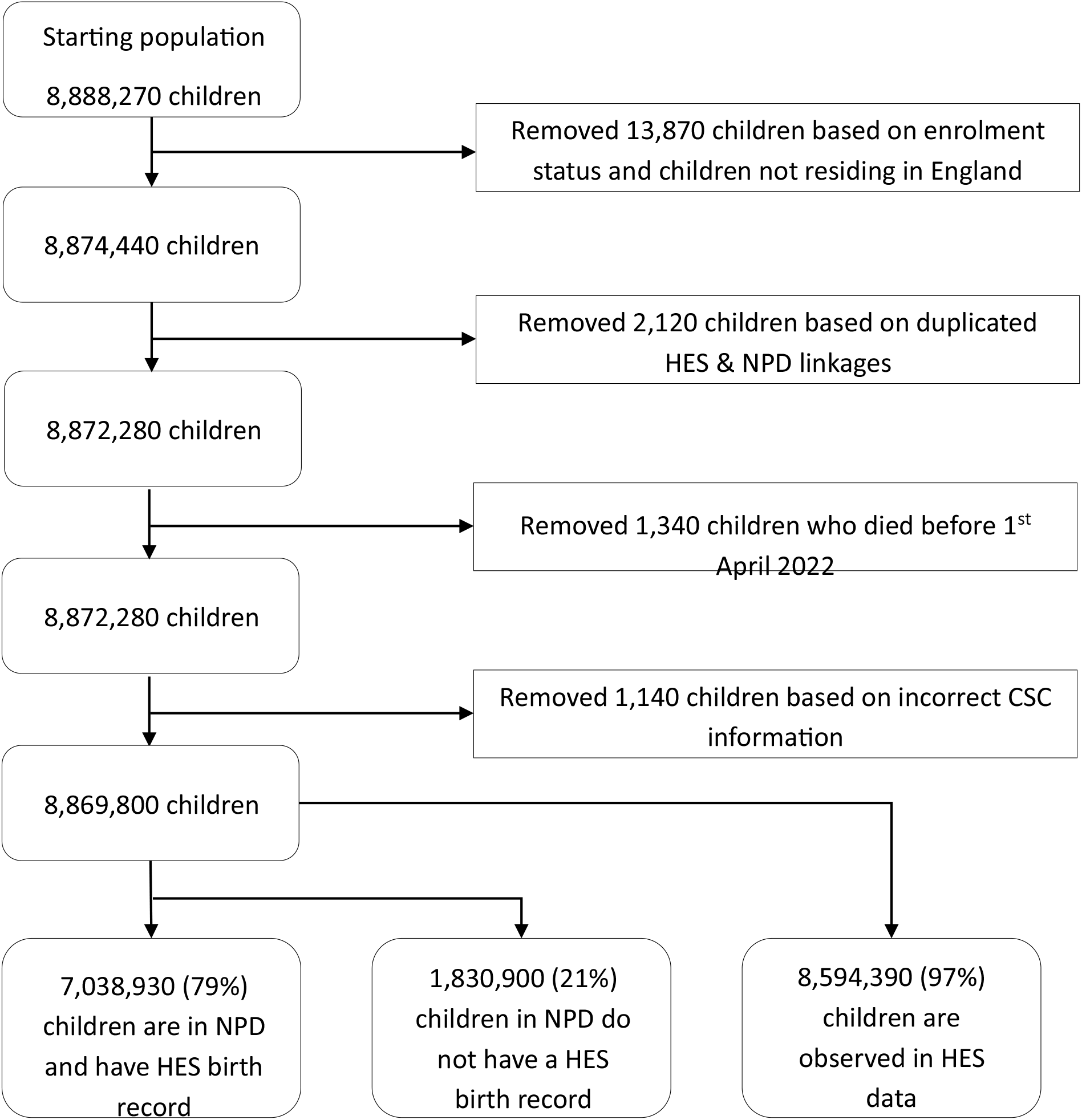
Population flow diagram. Note: abbreviations, NPD = National Pupil Database, HES = Hospital Episode Statistics

We initially identified 8,888,270 children based on anonymised unique pupil identifier. We removed children that do not reside in England as we may not be able to track hospital service utilisation of these children. The linkage between NPD and HES is imperfect, and as such we identified children with one HES ID linked to multiple unique pupil identifiers. For each case, we kept records which recorded the same birth month and year. Overall we omitted a total of 2,120 children due to linkage uncertainty. We observed 1,340 cases where a child dies in between the time between the school census date and 1^st^ April 2022. Finally, we removed 1,140 children who have conflicting age information between CSC data and the NPD.

This study selected a population of children that were based primarily on the NPD, as opposed to selecting individuals using birth cohorts. However, on limitation of using a birth cohort is not being able to track when children leave England and therefore are unable to use hospital services in HES financial year 2022/23. In total 79% of children in our cohort have a birth record in HES, where 97% of the population have been identified as using hospital services.

**Table A2:** Number and percentage of the population with at least one Hardelid condition by age and gender.

| Age | Children in the NPD |  |  |  | Children in the NPD and with a HES birth record |  |  |  |
| --- | --- | --- | --- | --- | --- | --- | --- | --- |
|  | Male |  | Female |  | Male |  | Female |  |
|  | Number | % of population | Number | % of population | Number | % of population | Number | % of population |
| 5 | 49,090 | 15.1 | 34,180 | 11.1 | 47,800 | 16.2 | 33,270 | 11.9 |
| 6 | 52,450 | 15.8 | 37,220 | 11.8 | 50,900 | 17.1 | 36,150 | 12.8 |
| 7 | 55,660 | 16.9 | 39,300 | 12.5 | 41,070 | 18.2 | 29,120 | 13.6 |
| 8 | 58,250 | 17.6 | 42,120 | 13.4 | 44,400 | 19.2 | 32,340 | 14.6 |
| 9 | 63,590 | 18.6 | 45,610 | 14 | 49,390 | 20.4 | 35,360 | 15.3 |
| 10 | 67,180 | 19.5 | 48,890 | 14.9 | 51,460 | 21.2 | 37,740 | 16.3 |
| 11 | 70,960 | 20.9 | 52,470 | 16.2 | 54,950 | 22.9 | 40,670 | 17.8 |
| 12 | 72,100 | 21.7 | 56,000 | 17.7 | 59,110 | 23.8 | 45,940 | 19.4 |
| 13 | 72,910 | 22.2 | 59,330 | 18.9 | 69,220 | 24.6 | 56,190 | 20.9 |
| 14 | 73,460 | 22.6 | 62,570 | 20.2 | 69,930 | 25.2 | 59,420 | 22.5 |
| 15 | 72,340 | 23.2 | 63,680 | 21.4 | 68,670 | 25.9 | 60,310 | 23.8 |
| 16 | 74,030 | 24.3 | 67,120 | 23.1 | 69,960 | 27.2 | 63,040 | 25.7 |
| 17 | 76,970 | 25.4 | 71,510 | 24.7 | 72,360 | 28.4 | 66,960 | 27.7 |
| 18 | 77,450 | 26.4 | 75,140 | 26.8 | 72,740 | 29.4 | 70,000 | 29.8 |

One limitation of using the sample of children that has a realistic opportunity to use hospital services in 2022/23 HES financial year, is that this sample will contain individuals that were not in England since birth and therefore not in the birth cohort. As the research conducted relies on administrative data, we would only identify needs such as health care needs (having CHC) if the child’s administrative hospital records record the diagnosis when using publicly funded hospital services in England. Therefore, when using data that is not also from the birth cohort, we may under count the number of children with a CHC need. Table A2 compares the numbers and percentage of the population that have a CHC recorded in their HES records. We find that children in the NPD as well as being in the birth record have higher population prevalence of CHC when compared to the sample of children in the NPD for both genders and across all ages. The lowest difference in prevalence is for females aged 5 at 0.8 percentage points, this percentage grows over time reaching three percentage points across 18 year olds (26.8% compares to 29.8%)

## Appendix B: Costing

### Number of children that have been costed by hospital service type

**Table B1:** Number of children that have used each of the costed hospital services.

|  | Population | Admitted Patient Care | Outpatient | Emergency Care | Critical Care |
| --- | --- | --- | --- | --- | --- |
| **Number of children** | 8,869,800 | 450,910 | 2,154,990 | 1,914,370 | 7,140 |
| **Percentage of population (%)** | 100 | 5.1 | 24.3 | 21.6 | 0.1 |

Table B1 shows the number of children that have costed service use among children aged five to 16 years. We find that in 2022/23 financial year, 24.3% of the population have used outpatient services contributing to over 8.6 million outpatient attendances. 21.6% of the population visited emergency department for either emergency or urgent care. 5.1% of children were admitted, and 0.1% of the population used critical care services.

### Proportion of hospital costs by service type and costing methods

**Table B2:**
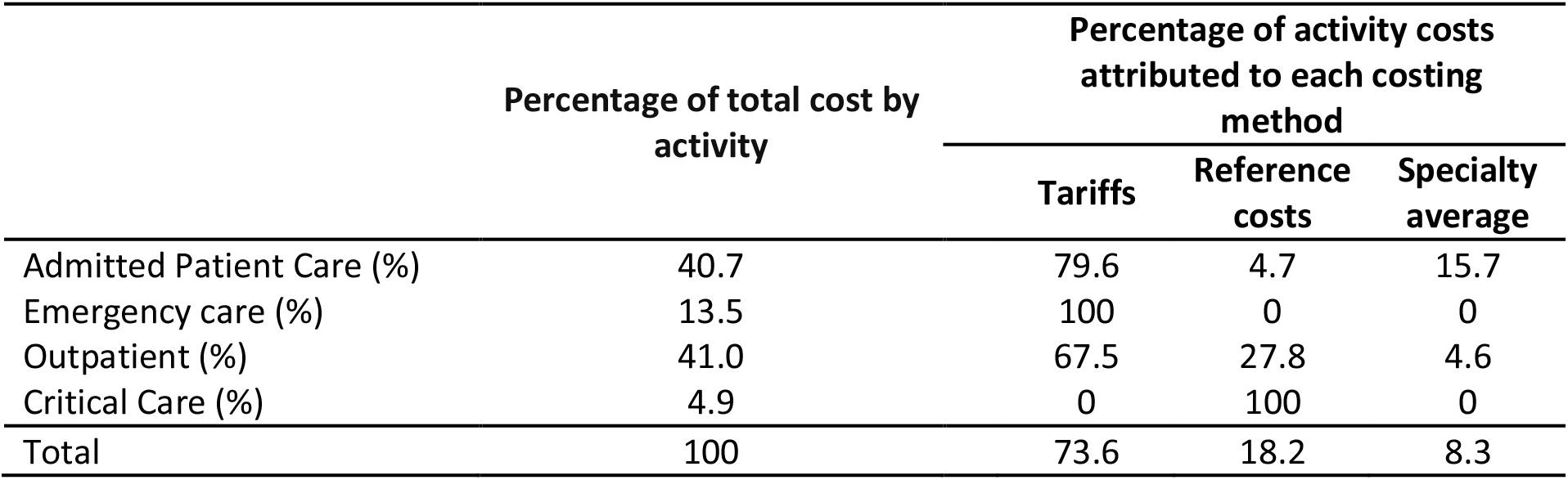
Percentage of total hospital costs contribution from hospital services in 2022/23 and proportion of costs attributed from each costing method.

Table B2 shows the percentages of total costs contributed from admitted patient care, outpatient services, and emergency department and from critical care in HES financial year 2022/23 for children ages between five and 16 as of 1^st^ April 2022. We find that Outpatient care contributes the largest percentage of overall costs (41.0%), whereas critical care the least (4.9%, despite only 0.1% of the population used critical care). 40.7% of total costs are attributed to inpatient services despite fewer children being admitted, this reflects higher costs of admissions compared to outpatient attendances.

Table B2 shows the proportion of costs that are attributable to each of the costing methods. All activity in emergency care is costed using HRG tariffs and critical care has been costed using only reference costs, where we were unable to cost critical care using tariffs. A total of 73.6% of all costs are costs using tariffs (79.6% all costs from admitted patient care and 67.5% of all outpatient services). 18.2% of total costs were from reference costs, mainly driven from outpatient services (27.8% of outpatient costs). Finally 8.3% of all costs are from using specialty averages (15.7% of inpatient costs and 4.6% of outpatient costs).

**Table B3:** Average costs in 2022/23 of children in eight categories by age and gender.

|  | None | CSC | SEND | CHC | CSC &<br>SEND | CSC &<br>CHC | SEND &<br>CHC | ALL |
| --- | --- | --- | --- | --- | --- | --- | --- | --- |
| Age | Males |  |  |  |  |  |  |  |
| 5 | 184 | 212 | 281 | 1005 | 323 | 1188 | 1944 | 2953 |
| 6 | 152 | 177 | 224 | 822 | 271 | 744 | 1440 | 1854 |
| 7 | 133 | 157 | 193 | 694 | 232 | 599 | 1193 | 2018 |
| 8 | 129 | 146 | 180 | 637 | 212 | 683 | 1116 | 1666 |
| 9 | 131 | 145 | 174 | 650 | 202 | 640 | 1018 | 1548 |
| 10 | 133 | 147 | 174 | 616 | 195 | 609 | 924 | 1553 |
| 11 | 142 | 162 | 177 | 680 | 202 | 710 | 976 | 1474 |
| 12 | 149 | 160 | 175 | 658 | 205 | 576 | 898 | 1316 |
| 13 | 163 | 177 | 180 | 691 | 197 | 659 | 867 | 1398 |
| 14 | 167 | 171 | 180 | 721 | 199 | 757 | 853 | 1521 |
| 15 | 158 | 175 | 170 | 736 | 183 | 695 | 926 | 1275 |
| 16 | 140 | 154 | 146 | 691 | 166 | 691 | 836 | 1329 |
| 17 | 131 | 136 | 134 | 668 | 147 | 743 | 825 | 1207 |
| 18 | 120 | 135 | 122 | 644 | 141 | 732 | 676 | 968 |
|  | Females |  |  |  |  |  |  |  |
| 5 | 165 | 214 | 282 | 1052 | 336 | 930 | 2522 | 3693 |
| 6 | 140 | 187 | 227 | 835 | 253 | 829 | 1962 | 3663 |
| 7 | 127 | 156 | 200 | 737 | 222 | 700 | 1688 | 2219 |
| 8 | 120 | 149 | 182 | 683 | 204 | 752 | 1512 | 2331 |
| 9 | 120 | 148 | 179 | 686 | 209 | 855 | 1259 | 2198 |
| 10 | 115 | 150 | 179 | 686 | 197 | 707 | 1228 | 1896 |
| 11 | 119 | 158 | 184 | 737 | 203 | 725 | 1186 | 1944 |
| 12 | 127 | 160 | 187 | 776 | 199 | 839 | 1280 | 2030 |
| 13 | 138 | 183 | 203 | 918 | 219 | 1092 | 1385 | 2075 |
| 14 | 151 | 189 | 205 | 1058 | 211 | 1145 | 1290 | 1899 |
| 15 | 153 | 181 | 186 | 1019 | 217 | 1196 | 1354 | 1814 |
| 16 | 141 | 184 | 172 | 935 | 202 | 1141 | 1162 | 1908 |
| 17 | 143 | 178 | 158 | 924 | 226 | 1286 | 1154 | 1855 |
| 18 | 148 | 179 | 149 | 867 | 209 | 1406 | 1091 | 1797 |

Note: Table shows average costs (£) in the eight mutually exclusive and exhaustive categories of children. These figures are shown in figure 1 in the main manuscript.

**Figure B1:**
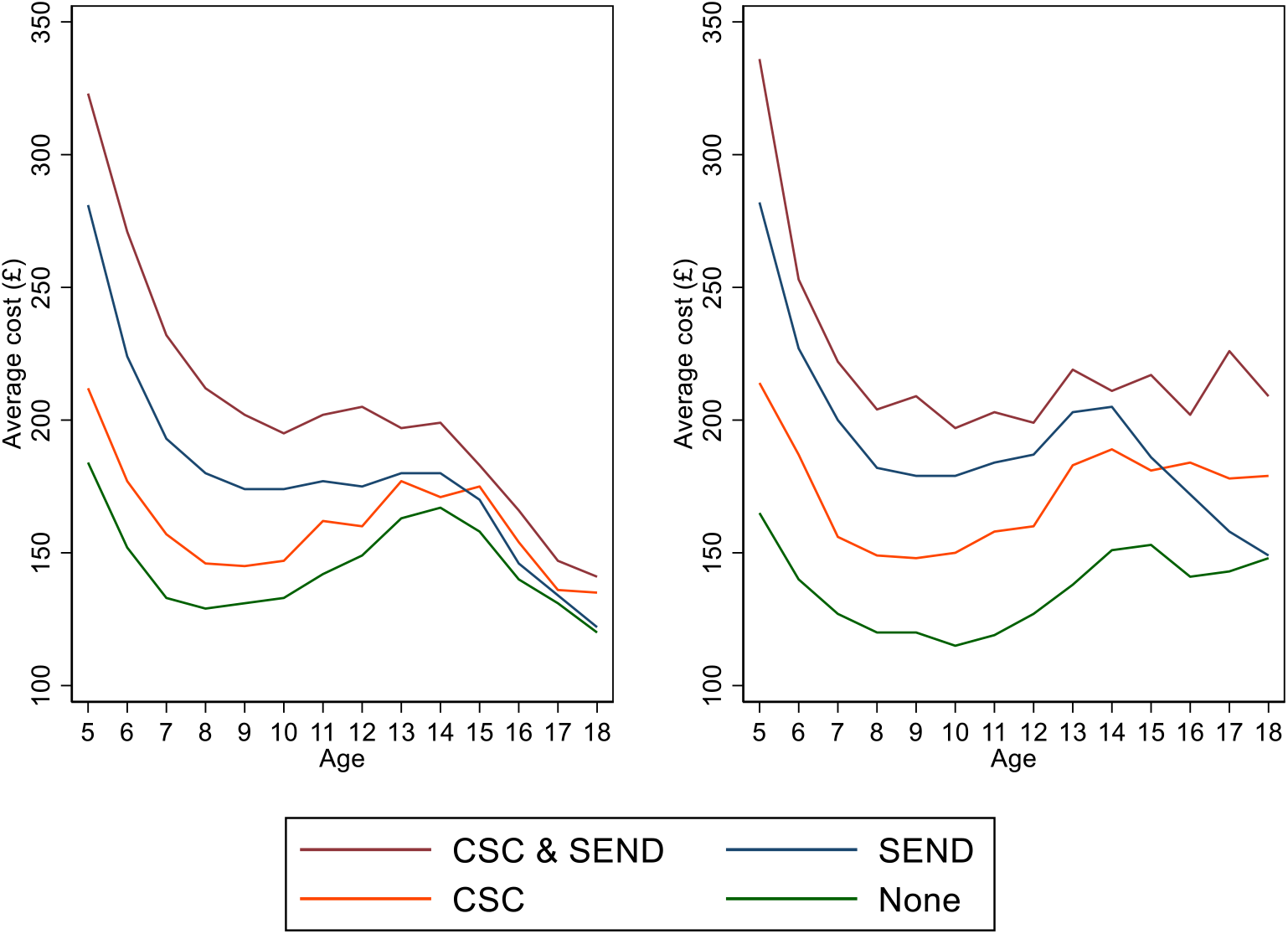
Average costs of the population in each needs category by age and gender without CHC.

Figure B1 shows average costs for four groups of children without a chronic health condition: without any additional support indicators; CSC only; SEND only: and CSC & SEND. We observe the lowest costs across all ages for both genders are individuals with no need categories, and highest average costs are for children with both CSC & SEND. We observe lowest costs for males are around 18 years of age whereas for females the lowest costs are observed before teenage years.

## Appendix C – Supplementary regression analysis

**Table C1:** Regression of hospital costs on needs indicators and their interactions for males, by age.

| Age | Main associations |  |  | Additional associations of two-way interactions |  |  | Additional association of three-way interaction | Regression constant term | Number of Children |
| --- | --- | --- | --- | --- | --- | --- | --- | --- | --- |
|  | CSC | SEND | CHC | CSC&SEND | CSC&CHC | SEND&CHC |  |  |  |
| 5 | 28<br>[13,42] | 97<br>[92,103] | 821<br>[761,882] | 14<br>[-15,44] | 155<br>[-428,738] | 841<br>[694,989] | 811<br>[-91,1714] | 184<br>[182,186] | 324270 |
| 6 | 25<br>[13,36] | 72<br>[67,77] | 670<br>[622,718] | 22<br>[2,41] | -102<br>[-276,72] | 546<br>[453,639] | 470<br>[148,793] | 152<br>[150,154] | 331390 |
| 7 | 24<br>[15,33] | 60<br>[55,64] | 561<br>[511,611] | 15<br>[-0,31] | -119<br>[-235,-3] | 439<br>[343,534] | 905<br>[576,1234] | 133<br>[131,135] | 329000 |
| 8 | 17<br>[9,25] | 51<br>[47,55] | 508<br>[459,558] | 15<br>[2,29] | 29<br>[-90,148] | 427<br>[343,511] | 490<br>[272,708] | 129<br>[127,131] | 330850 |
| 9 | 14<br>[6,22] | 43<br>[38,47] | 519<br>[476,563] | 14<br>[2,27] | -24<br>[-142,93] | 325<br>[239,410] | 526<br>[280,772] | 131<br>[129,133] | 341830 |
| 10 | 14<br>[7,21] | 41<br>[37,46] | 483<br>[446,520] | 7<br>[-5,18] | -21<br>[-113,70] | 266<br>[191,342] | 630<br>[430,830] | 133<br>[131,135] | 344010 |
| 11 | 20<br>[12,28] | 35<br>[28,42] | 538<br>[482,594] | 5<br>[-8,18] | 10<br>[-242,262] | 261<br>[162,359] | 464<br>[147,780] | 142<br>[139,145] | 340240 |
| 12 | 11<br>[4,19] | 26<br>[21,30] | 509<br>[465,553] | 19<br>[7,30] | -94<br>[-173,-14] | 214<br>[138,290] | 482<br>[326,639] | 149<br>[147,152] | 332730 |
| 13 | 14<br>[4,24] | 17<br>[12,22] | 528<br>[487,570] | 3<br>[-10,15] | -46<br>[-132,40] | 158<br>[99,218] | 560<br>[395,726] | 163<br>[161,166] | 327910 |
| 14 | 4<br>[-4,13] | 13<br>[8,18] | 554<br>[508,599] | 15<br>[3,26] | 32<br>[-88,152] | 119<br>[55,183] | 617<br>[421,812] | 167<br>[164,170] | 325080 |
| 15 | 17<br>[8,26] | 12<br>[7,17] | 578<br>[535,622] | -4<br>[-16,8] | -58<br>[-165,49] | 178<br>[81,275] | 394<br>[220,567] | 158<br>[155,161] | 311820 |
| 16 | 14<br>[6,23] | 6<br>[0,11] | 551<br>[502,600] | 6<br>[-6,18] | -14<br>[-102,74] | 140<br>[68,212] | 487<br>[321,652] | 140<br>[137,143] | 304720 |
| 17 | 5<br>[-4,13] | 3<br>[-2,8] | 537<br>[500,573] | 8<br>[-3,20] | 71<br>[-19,162] | 155<br>[80,229] | 297<br>[138,457] | 131<br>[128,134] | 302710 |
| 18 | 15<br>[6,23] | 2<br>[-3,7] | 524<br>[487,561] | 4<br>[-8,16] | 74<br>[-38,186] | 29<br>[-40,98] | 200<br>[55,345] | 120<br>[117,123] | 292780 |

Notes: Coefficients are from age and gender stratified linear regressions including all eight terms (constant term, three main associations, three two-way interactions, and one three-way interaction). 95% confidence intervals shown around each estimate. CSC is an indicator for whether the child has received social care support. SEND is an indicator for whether the child has a special educational need or disability or an educational health care plan. CHC is an indicator for whether the child has a chronic health condition. ALL represents children with CSC, SEND and CHC.

**Table C2:**
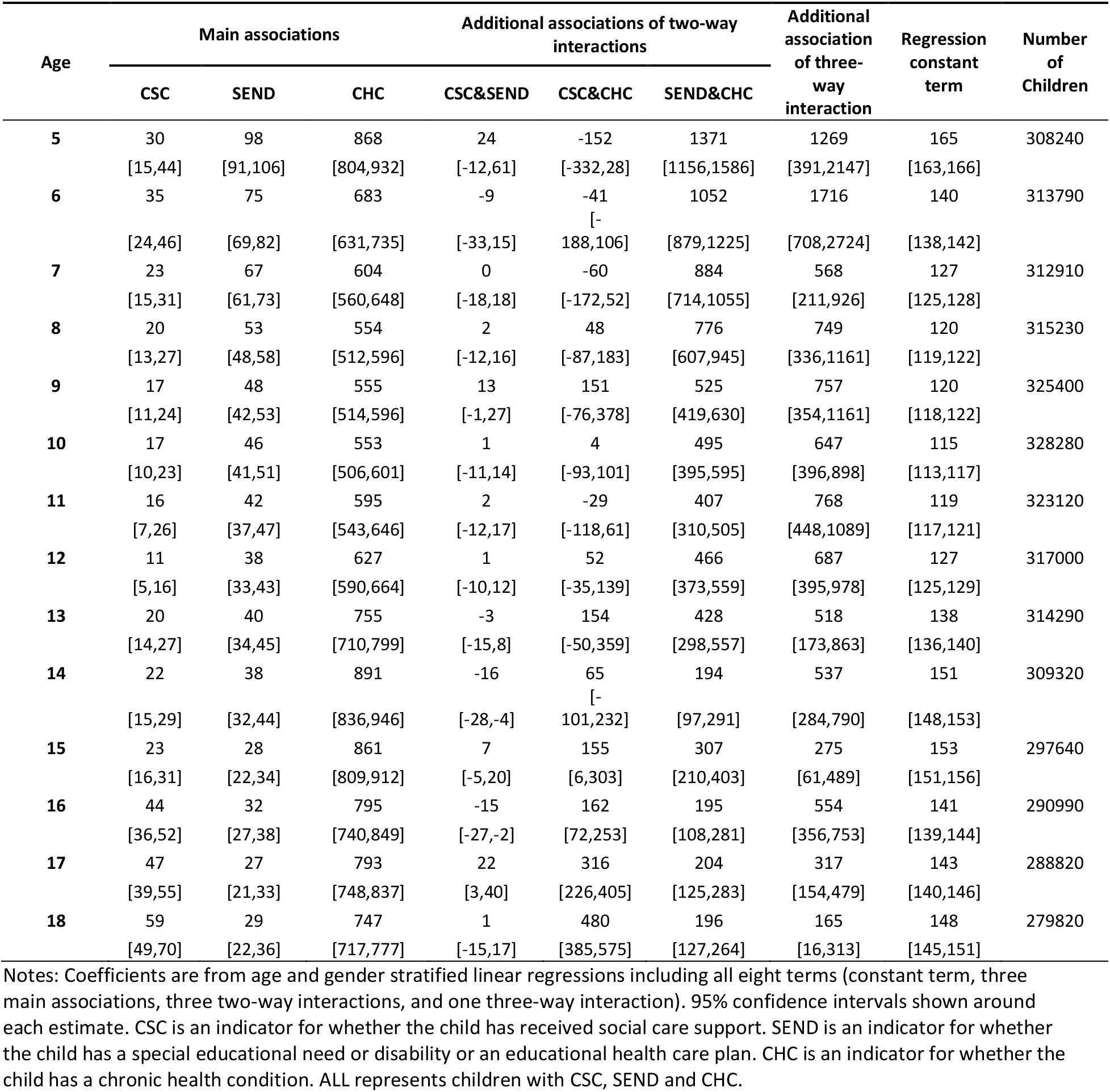
Regression of hospital costs on needs indicators and their interactions for females, by age.

**Table C3:**
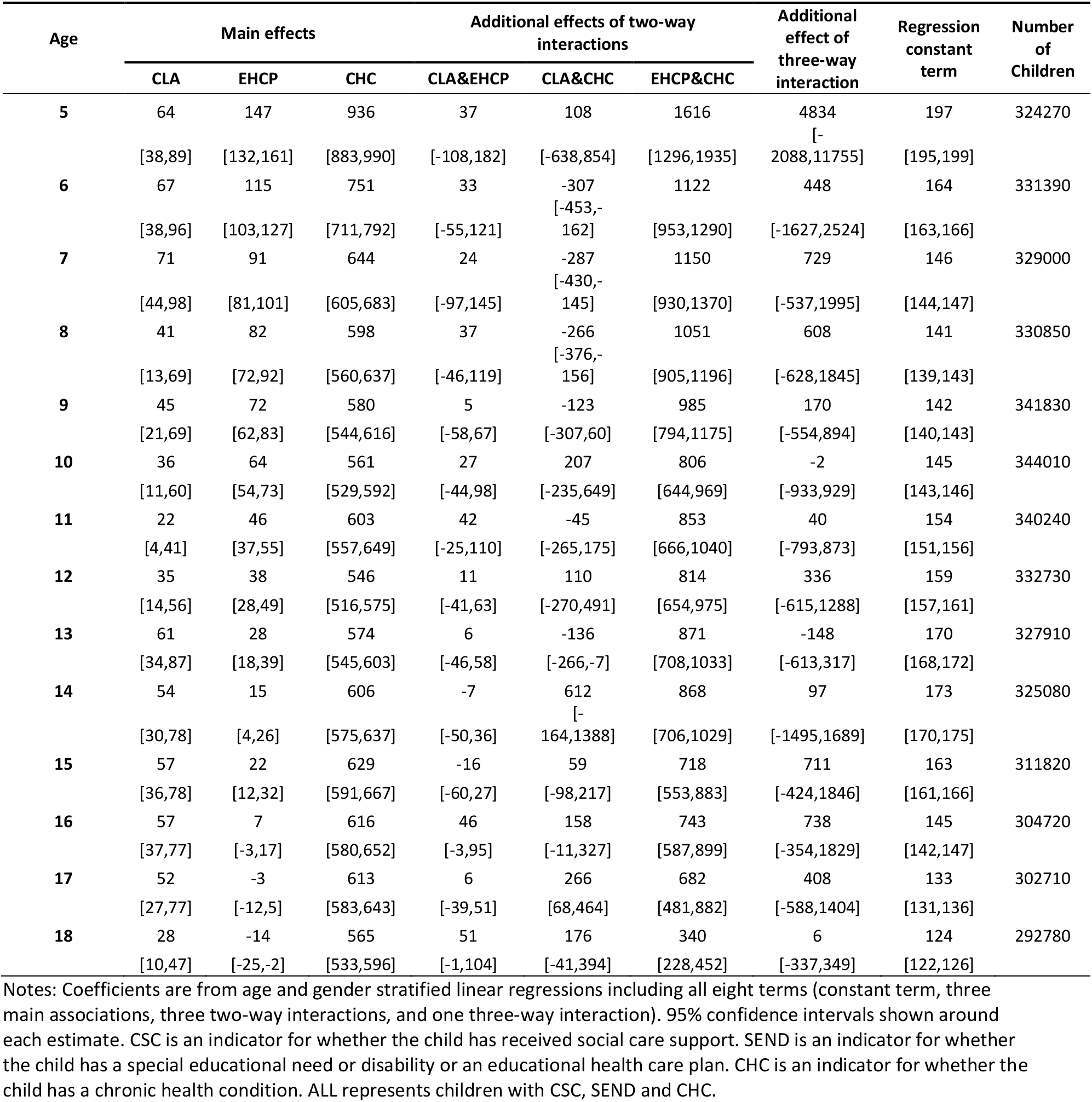
Regression of hospital costs on higher levels of needs indicators (education and healthcare plan - EHCP and child looked after - CLA) and their interactions for males, by age.

**Table C4:** Regression of hospital costs on higher levels of needs indicators (education and healthcare plan - EHCP and child looked after - CLA) and their interactions for females, by age.

| Age | Main effects |  |  | Additional effects of two-way interactions |  |  | Additional effect of three-way interaction | Regression constant term | Number of Children |
| --- | --- | --- | --- | --- | --- | --- | --- | --- | --- |
|  | CLA | EHCP | CHC | CLA&EHCP | CLA&CHC | EHCP&CHC |  |  |  |
| 5 | 64 | 147 | 936 | 37 | 108 | 1616 | 4834 | 197 | 324270 |
|  | [38,89] | [132,161] | [883,990] | [-108,182] | [-638,854] | [1296,1935] | [-2088,11755] | [195,199] |  |
| 6 | 67 | 115 | 751 | 33 | -307 | 1122 | 448 | 164 | 331390 |
|  | [38,96] | [103,127] | [711,792] | [-55,121] | [-453,-162] | [953,1290] | [-1627,2524] | [163,166] |  |
| 7 | 71 | 91 | 644 | 24 | -287 | 1150 | 729 | 146 | 329000 |
|  | [44,98] | [81,101] | [605,683] | [-97,145] | [-430,-145] | [930,1370] | [-537,1995] | [144,147] |  |
| 8 | 41 | 82 | 598 | 37 | -266 | 1051 | 608 | 141 | 330850 |
|  | [13,69] | [72,92] | [560,637] | [-46,119] | [-376,-156] | [905,1196] | [-628,1845] | [139,143] |  |
| 9 | 45 | 72 | 580 | 5 | -123 | 985 | 170 | 142 | 341830 |
|  | [21,69] | [62,83] | [544,616] | [-58,67] | [-307,60] | [794,1175] | [-554,894] | [140,143] |  |
| 10 | 36 | 64 | 561 | 27 | 207 | 806 | -2 | 145 | 344010 |
|  | [11,60] | [54,73] | [529,592] | [-44,98] | [-235,649] | [644,969] | [-933,929] | [143,146] |  |
| 11 | 22 | 46 | 603 | 42 | -45 | 853 | 40 | 154 | 340240 |
|  | [4,41] | [37,55] | [557,649] | [-25,110] | [-265,175] | [666,1040] | [-793,873] | [151,156] |  |
| 12 | 35 | 38 | 546 | 11 | 110 | 814 | 336 | 159 | 332730 |
|  | [14,56] | [28,49] | [516,575] | [-41,63] | [-270,491] | [654,975] | [-615,1288] | [157,161] |  |
| 13 | 61 | 28 | 574 | 6 | -136 | 871 | -148 | 170 | 327910 |
|  | [34,87] | [18,39] | [545,603] | [-46,58] | [-266,-7] | [708,1033] | [-613,317] | [168,172] |  |
| 14 | 54 | 15 | 606 | -7 | 612 | 868 | 97 | 173 | 325080 |
|  | [30,78] | [4,26] | [575,637] | [-50,36] | [-164,1388] | [706,1029] | [-1495,1689] | [170,175] |  |
| 15 | 57 | 22 | 629 | -16 | 59 | 718 | 711 | 163 | 311820 |
|  | [36,78] | [12,32] | [591,667] | [-60,27] | [-98,217] | [553,883] | [-424,1846] | [161,166] |  |
| 16 | 57 | 7 | 616 | 46 | 158 | 743 | 738 | 145 | 304720 |
|  | [37,77] | [-3,17] | [580,652] | [-3,95] | [-11,327] | [587,899] | [-354,1829] | [142,147] |  |
| 17 | 52 | -3 | 613 | 6 | 266 | 682 | 408 | 133 | 302710 |
|  | [27,77] | [-12,5] | [583,643] | [-39,51] | [68,464] | [481,882] | [-588,1404] | [131,136] |  |
| 18 | 28 | -14 | 565 | 51 | 176 | 340 | 6 | 124 | 292780 |
|  | [10,47] | [-25,-2] | [533,596] | [-1,104] | [-41,394] | [228,452] | [-337,349] | [122,126] |  |

Notes: Coefficients are from age and gender stratified linear regressions including all eight terms (constant term, three main associations, three two-way interactions, and one three-way interaction). 95% confidence intervals shown around each estimate. CSC is an indicator for whether the child has received social care support. SEND is an indicator for whether the child has a special educational need or disability or an educational health care plan. CHC is an indicator for whether the child has a chronic health condition. ALL represents children with CSC, SEND and CHC.

**Table C5:** Regression of hospital costs on needs indicators and their interactions for males, by age, restricting SEND recording to previous 4 years.

| Age | Main effects |  |  | Additional effects of two-way interactions |  |  | Additional effect of three-way interaction | Regression constant term | Number of Children |
| --- | --- | --- | --- | --- | --- | --- | --- | --- | --- |
|  | CSC | SEND | CHC | CSC&SEND | CSC&CHC | SEND&CHC |  |  |  |
| 5 | 28 | 97 | 821 | 14 | 155 | 841 | 811 | 184 | 324270 |
|  | [13,42] | [92,103] | [761,882] | [-15,44] | [-428,738] | [694,989] | [-91,1714] | [182,186] | 0 |
| 6 | 25 | 72 | 670 | 22 | -102 | 546 | 470 | 152 | 331390 |
|  | [13,36] | [67,77] | [622,718] | [2,41] | [-276,72] | [453,639] | [148,793] | [150,154] |  |
| 7 | 24 | 60 | 561 | 15 | -119 | 439 | 905 | 133 | 329000 |
|  | [15,33] | [55,64] | [511,611] | [-0,31] | [-235,-3] | [343,534] | [576,1234] | [131,135] | 0 |
| 8 | 17 | 51 | 508 | 15 | 29 | 427 | 490 | 129 | 330850 |
|  | [9,25] | [47,55] | [459,558] | [2,29] | [-90,148] | [343,511] | [272,708] | [127,131] | 0 |
| 9 | 13 | 44 | 519 | 15 | -29 | 334 | 534 | 131 | 341830 |
|  | [6,21] | [39,48] | [476,562] | [3,27] | [-144,85] | [248,421] | [287,782] | [129,133] |  |
| 10 | 14 | 46 | 486 | 5 | -5 | 275 | 619 | 133 | 344010 |
|  | [7,21] | [41,50] | [450,522] | [-6,17] | [-100,90] | [197,352] | [412,825] | [131,135] |  |
| 11 | 21 | 40 | 548 | 2 | -19 | 264 | 528 | 142 | 340240 |
|  | [13,29] | [32,48] | [488,609] | [-12,15] | [-244,207] | [166,362] | [227,828] | [140,145] |  |
| 12 | 9 | 30 | 508 | 26 | -85 | 251 | 486 | 150 | 332730 |
|  | [2,16] | [25,35] | [467,549] | [14,38] | [-161,-10] | [170,333] | [322,651] | [147,152] |  |
| 13 | 12 | 26 | 523 | 5 | -40 | 211 | 589 | 163 | 327910 |
|  | [3,20] | [20,31] | [485,561] | [-8,17] | [-117,37] | [150,272] | [414,765] | [160,166] |  |
| 14 | 6 | 22 | 548 | 12 | 52 | 174 | 650 | 166 | 325080 |
|  | [-2,13] | [16,28] | [509,586] | [-0,24] | [-53,157] | [106,243] | [444,856] | [164,169] |  |
| 15 | 9 | 18 | 586 | 11 | -42 | 244 | 418 | 159 | 311820 |
|  | [2,16] | [12,25] | [540,633] | [-2,23] | [-128,44] | [124,364] | [228,608] | [156,161] |  |
| 16 | 10 | 9 | 539 | 20 | -18 | 277 | 559 | 141 | 304720 |
|  | [3,17] | [3,16] | [501,578] | [8,33] | [-82,45] | [187,368] | [361,756] | [138,143] |  |
| 17 | 7 | 9 | 591 | 5 | 55 | 74 | 479 | 131 | 302710 |
|  | [1,14] | [3,16] | [549,633] | [-7,17] | [-27,138] | [-10,158] | [292,666] | [128,133] |  |
| 18 | 14 | 5 | 523 | 7 | 99 | 66 | 222 | 120 | 292780 |
|  | [8,21] | [-1,12] | [488,557] | [-6,21] | [12,187] | [-33,165] | [67,378] | [117,122] |  |

Notes: Coefficients are from age and gender stratified linear regressions including all eight terms (constant term, three main associations, three two-way interactions, and one three-way interaction). 95% confidence intervals shown around each estimate. CSC is an indicator for whether the child has received social care support. SEND is an indicator for whether the child has a special educational need or disability or an educational health care plan. CHC is an indicator for whether the child has a chronic health condition. ALL represents children with CSC, SEND and CHC. Children are classed as having SEND if they received SEND within the prior 4 years.

**Table C6:** Regression of hospital costs on needs indicators and their interactions for females, by age, restricting SEND recording to previous 4 years.

| Age | Main effects |  |  | Additional effects of two-way interactions |  |  | Additional effect of three-way interaction | Regression constant term | Number of Children |
| --- | --- | --- | --- | --- | --- | --- | --- | --- | --- |
|  | CSC | SEND | CHC | CSC&SEND | CSC&CHC | SEND&CHC |  |  |  |
| 5 | 30 | 98 | 868 | 24 | -152 | 1371 | 1269 | 165 | 308240 |
|  | [15,44] | [91,106] | [804,932] | [-12,61] | [-332,28] | [1156,1586] | [391,2147] | [163,166] | 0 |
| 6 | 35 | 75 | 683 | -9 | -41 | 1052 | 1716 | 140 | 313790 |
|  | [24,46] | [69,82] | [631,735] | [-33,15] | [-188,106] | [879,1225] | [708,2724] | [138,142] |  |
| 7 | 23 | 67 | 604 | 0 | -60 | 884 | 568 | 127 | 312910 |
|  | [15,31] | [61,73] | [560,648] | [-18,18] | [-172,52] | [714,1055] | [211,926] | [125,128] | 0 |
| 8 | 20 | 53 | 554 | 2 | 48 | 776 | 749 | 120 | 315230 |
|  | [13,27] | [48,58] | [512,596] | [-12,16] | [-87,183] | [607,945] | [336,1161] | [119,122] | 0 |
| 9 | 17 | 49 | 555 | 13 | 148 | 540 | 765 | 120 | 325400 |
|  | [11,24] | [44,55] | [515,595] | [-1,27] | [-75,372] | [433,648] | [360,1171] | [118,122] |  |
| 10 | 16 | 47 | 560 | 4 | -9 | 503 | 698 | 116 | 328280 |
|  | [10,22] | [42,52] | [512,608] | [-8,17] | [-103,85] | [403,603] | [440,956] | [114,117] |  |
| 11 | 17 | 48 | 594 | 1 | -6 | 458 | 767 | 119 | 323120 |
|  | [8,26] | [43,54] | [545,644] | [-14,16] | [-105,92] | [354,562] | [423,1110] | [117,121] |  |
| 12 | 11 | 46 | 641 | 0 | 34 | 491 | 806 | 127 | 317000 |
|  | [5,16] | [40,51] | [602,679] | [-12,12] | [-50,117] | [394,588] | [486,1125] | [125,129] |  |
| 13 | 19 | 50 | 749 | -1 | 145 | 557 | 574 | 139 | 314290 |
|  | [13,25] | [44,56] | [707,791] | [-14,11] | [-30,321] | [402,712] | [203,944] | [137,141] |  |
| 14 | 19 | 55 | 880 | -13 | 43 | 297 | 662 | 151 | 309320 |
|  | [12,25] | [47,62] | [829,930] | [-27,0] | [-95,181] | [185,409] | [392,932] | [148,153] |  |
| 15 | 25 | 50 | 868 | 2 | 137 | 414 | 382 | 153 | 297640 |
|  | [19,32] | [42,58] | [819,917] | [-13,17] | [18,255] | [304,523] | [153,610] | [151,156] |  |
| 16 | 43 | 53 | 796 | -21 | 196 | 317 | 666 | 143 | 290990 |
|  | [36,49] | [45,62] | [748,843] | [-36,-5] | [118,275] | [209,424] | [406,926] | [140,145] |  |
| 17 | 51 | 40 | 794 | 33 | 359 | 379 | 300 | 145 | 288820 |
|  | [44,58] | [31,49] | [755,833] | [1,65] | [287,431] | [271,488] | [83,517] | [143,148] |  |
| 18 | 61 | 35 | 772 | 8 | 517 | 263 | 167 | 152 | 279820 |
|  | [53,70] | [25,45] | [745,799] | [-14,31] | [441,593] | [148,378] | [-29,363] | [149,154] |  |

Notes: Coefficients are from age and gender stratified linear regressions including all eight terms (constant term, three main associations, three two-way interactions, and one three-way interaction). 95% confidence intervals shown around each estimate. CSC is an indicator for whether the child has received social care support. SEND is an indicator for whether the child has a special educational need or disability or an educational health care plan. CHC is an indicator for whether the child has a chronic health condition. ALL represents children with CSC, SEND and CHC. Children are classed as having SEND if they received SEND within the prior 4 years.

